# The impact of public-initiated COVID-19 risk communication on individual NPI practices

**DOI:** 10.1101/2023.03.07.23286938

**Authors:** Yifokire Tefera, Abera Kumie, Damen Hailemariam, Samson Wakuma, Teferi Abegaz, Shibabaw Yirsaw

**Affiliations:** Department of Preventive Medicine, School of Public Health, College of Health Sciences, Addis Ababa University, Addis Ababa, Ethiopia

## Abstract

**Background:** Non-Pharmaceutical Interventions (NPI) are the most widely recognized public health measures practiced globally to prevent the spread of Covid-19 transmission. The effectiveness of NPIs is dependent on the type, a combination of applied interventions, and the level of compliance of the NPIs. The expected outcome of behavioral practices varies relative to the behavioral intervention duration.

**Objectives:** This study aimed to assess the trend of community compliance to NPIs and with its level of variation with the place of residence and sociodemographic characteristics of people.

**Methods:** A weekly non-participatory field survey on an individual’s NPI practice was observed from October 2020 to July 2021, for a total of 39 weeks. The survey covered all the regions; 14 regional and capital cities. Data collection for the 3 NPI behaviors (mask use, hand hygiene, and physical distance was and managed weekly at eight public service locations using the Open Data Kit (ODK) tool.

**Results:** More than 180,000 individuals were observed for their NPI practice; on average 5,000 observations in a week. About 43% of the observation was made from Addis Ababa, 56% were male participants and the middle age group (18-50 years) accounts for 75%. The overall level of NPI compliance had a peak around the 26^th^ – 30^th^ weeks then decline the rest of the weeks. Respiratory hygiene had the highest compliance whereas hand hygiene had the least 41% and 4%, respectively. There was a significant difference between capital city and regional city residents by their level of NPI compliance. Females comply more than males, and individuals had increased NPI compliance while at the bank service and workplaces compared to while in the transport services at p<0.001.

**Conclusion:** The overall community compliance with NPI practice showed a declining trend in Ethiopia but increased compliance was also observed following the implementation of government-initiated public measures. Therefore, public-initiated risk communication and public advocacy programs for the prevention of Covid-19 should be strengthened.

## Introduction

Individual level Non-Pharmaceutical Intervention (NPI) is part of the bigger public health and social measures (PHSM) of the World Health Organization (WHO) to prevent the spread of Covid-19 transmission. These behaviors include putting on a mask, having physical distancing, and using proper hand hygiene [1]. In the early stages of Covid-19, governments from all over the world started a variety of PHSMs that can be implemented by individuals, institutions, communities, and local and national government bodies due to the lack of clear and proven medical treatments for Covid-19 and the paucity of global level available scientific evidence [1-4]. Countries and governments were under pressure as disease incidence rose.

Individual behavioral practices are advised as vital to halt the development of COVID-19, although countries have employed a variety of context-based, diverse PHSM interventions [5]. Few studies attempted to assess the efficacy of various NPI practices adopted by governments of various nations [2, 3]. Haug and colleagues [2] gathered thousands of implemented NPIs from 79 different countries and discovered that no one best NPI can stop the spread of Covid-19. Instead, the researchers found several combinations of interventions that can drastically reduce the transmission rate. Curfews, limitations, lockdowns, as well as limiting and restricting locations for public meetings, were shown to be the most effective NPIs in this study [2]. In a comparable cross-country study,[3] assessed the efficacy of eight NPI interventions related to case identification, environmental measures, health care, and public health capacity, resource allocation, risk communication, social distancing, travel restriction, and returning to normal life. This study demonstrated that risk communication had the greatest impact on the population as a whole. The authors emphasized that risk communication strategies, such as providing general information about Covid-19 or using a face mask, were less likely to enforce any particular behavior on individuals but had a significant impact on the public.

The WHO created the “Covid-19 Global Risk Communication and Community Engagement Strategy” after considering risk communication to be a potent tool for altering people’s behavior and willingness to adhere to public health measures [6]. The ability to compare the impact of interventions was limited since many NPIs were introduced at the same time or at separate times [2, 5, 7]. Sharma and colleagues mathematical model [4, 7] highlighted the importance of local context and suggested a dozen assumptions be taken into account for a reliable estimation of the effect of NPIs [4, 7]. However, several studies concurred and suggested that combined NPIs were the most efficient way to limit the spread of Covid-19 [2-5, 8].

The first Covid-19 case was found in Ethiopia in the second week of March 2020, and as of the most recent report (October 18, 2022), there were about 500,000 instances of infection [9]. Since the introduction of the infection, the federal government has put several NPI measures into effect across the nation, including the implementation of the provisions of a State of Emergency, such as the closing of the international land border, the cancellation, closure, and restriction of public gatherings, school uniforms, public awareness campaigns, personal protective measures, case detection, isolation, and quarantine, among other things. Regional governments had also placed the various implementation strategies within the context of local and regional capabilities [1, 10]. The Federal Ministry of Health’s (FMoH) Ethiopian Public Health Institute (EPHI) takes the lead in coordination with regional partners and governments [10, 11].

The three NPI measures—mask use, physical separation, and hand hygiene—were the Covid-19 interventions that were used the most frequently on an individual basis. In the second week of June 2020, [12] investigated these personal safety precautions among Addis Ababa city government employees. They found that more than 90% of participants used face masks, washed their hands, and maintained physical distancing [12]. A population of 12,056 residents of Addis Ababa city participated in a weekly NPI monitoring cross-sectional non-participatory observation from April to June 2020 for 10 uninterrupted weeks. The study found an increase in proper hand hygiene from the baseline of 24% to 33%, proper physical distance from 34% to 43%, and mask use from 24% to 77% in week 10 [11]. This study showed the peak of practicing proper respiratory hygiene took 6 weeks after the baseline within the context of practicing the measures of State Emergency [13]. Another study in the country, Bule Hora Town West Guji Zone, during the last weeks of September 2020 found that 38% of the participants had good social distancing practices [14].

We developed a research question associated with the level, and trend of NPIs after the lifting of the actions of the State of Emergency: what are the level and likely trends in the vacuum of public-supported NPI intervention? The current study focuses on determining the level, and the trend of the weekly changes in the three individual-level NPI behaviors. This is an urban population and time-expanded extension of the weekly NPI monitoring non-participatory observational research [11].

## Methods

### Study design, study area, setting, and period

This cross-sectional study design tracked people’s weekly NPI behaviors in several public places throughout the urban centers of Ethiopia. The NPI techniques of people monitoring were first implemented in Addis Ababa in April 2020, and they have since spread throughout the entire nation, encompassing all regional capital cities. Before 2020, there were nine regional states, but the Federal Government of Ethiopia now has 11 regional states and 2 chartered cities. Two new areas, Sidama and South West Ethiopia Regions, were just established in June 2020. Weekly monitoring data from 15 cities, including Addis Abeba, Bahir Dar, Gondar, Adama, Hawasa, Asosa, Gambella, Diredewa, Harar, Hosana, Jigjiga, Jimma, Semera, Mekele, and Welayta Sodo, were obtained for the current study.

Ethiopia’s overall population is expected to be 105,166 000 in 2022, according to CSA projections [15]. The data collection period spanned 39 weeks, from October 4, 2020, to July 4, 2021, with a 2-week interruption in the middle (27^th^ and 28^th^ weeks).

The three NPI behaviors (mask use, hand hygiene, and physical distance) were observed weekly at eight public service locations, including places of worship, medical institutions, markets, banks, public transportation hubs, restaurants, and workplaces. These sites assumed an increase in COVID-19 transmission when individuals were taking public services, such as marketing, accessing transport services, and attending churches.

### Source population

About 23,880, 000 Ethiopian urban inhabitants served as the study’s source population [15]. Around 7,333,908 people resided in the study’s sample towns in total. The location of each town is indicated in Figure 1.

**Figure 1:**
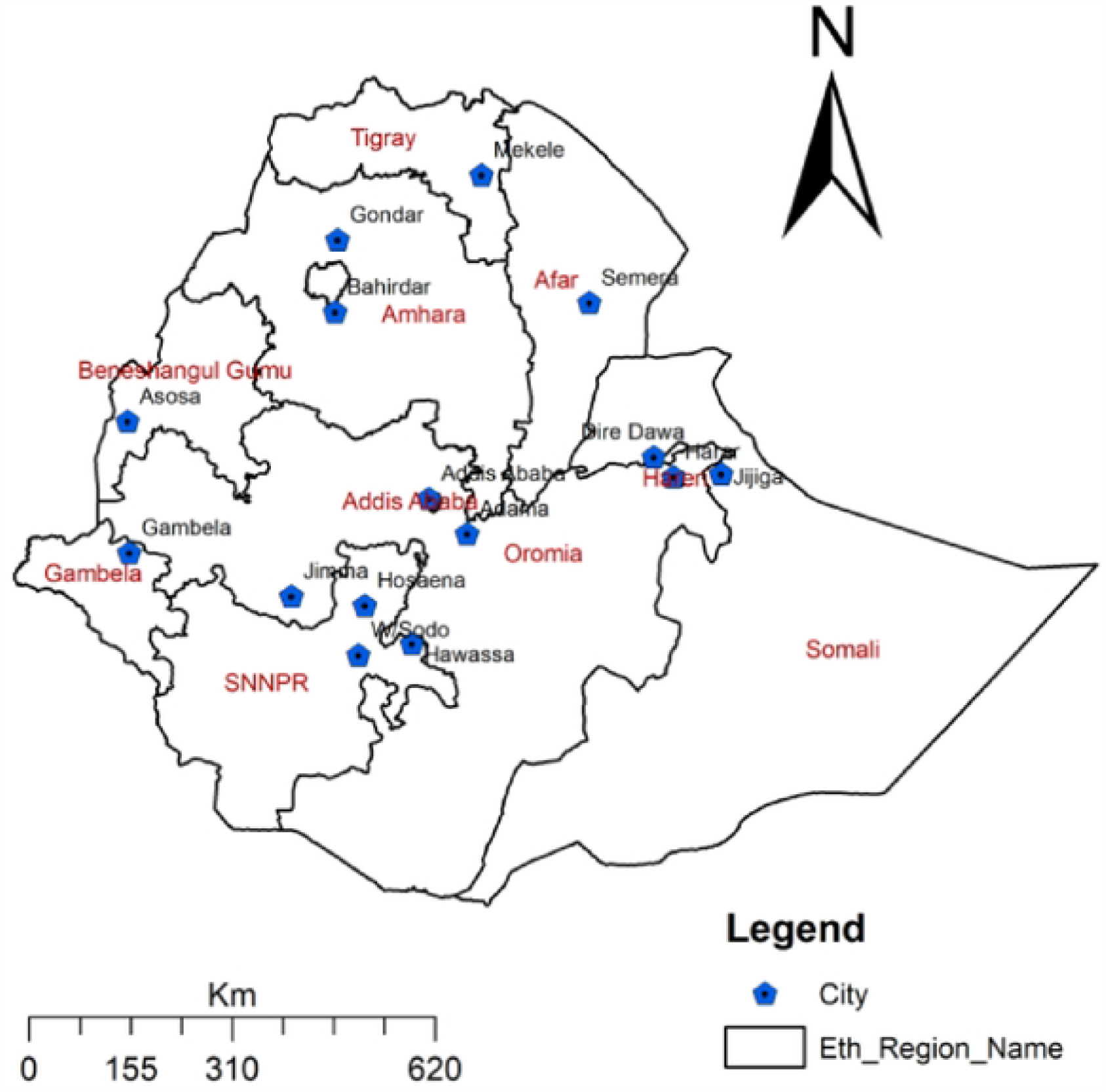
Location of COVID-19 NPls MonitoringSites by City, Ethiopia, October/2020-July 2021.

### Monitoring protocol, Data collection tools, and Data collection procedure

This study is an extension of the Covid-19 monitoring in Ethiopia, hence monitoring protocol, data collection tools, and data collection procedure was similar to the description mentioned in the previously published report [11].

### Data management and analysis

For data management and analysis, the ODK server’s raw data were obtained in excel format and exported to SPSS V 26 for data cleaning and analysis. All regional town observation data were combined to compare with Addis Ababa because that city’s population made up 51% of the study’s total population. Tables, figures, and descriptive statistics were used to present the data. Line and bar graphs were used to display the weekly trend, variations by service facilities, sex, and age group, and appropriate mask use, appropriate hand hygiene, and appropriate physical distancing practices. Using the Chi-square test, the level of NPI practices in Addis Ababa and nearby regional cities was examined.

### Ethical considerations

This study had been granted ethical approval from the Institutional Review Board (IRB) of the College of Health Sciences at Addis Ababa University. The detailed procedure was presented in the previous publication [11]. Data collection was anonymous and the observers acted similarly to study subjects in observation sites when collecting data.

## Results

### Study participants

More than 180,000 people were weekly observed for their NPI practice for a period of 39 weeks, although the observation was halted for two weeks. On average 5,000 weekly observations have been made from Addis Ababa and major regional cities. The proportion of observation from the capital city, Addis Ababa was 43% and from major regional cities 57%. Males’ participation in the observation had a slightly higher proportion, 56%, compared to females, 44%. The middle age group, 18-50 years were the biggest proportion accounts for 75% followed by the highest age group greater than 50 years accounts 14% (Table 1)

**Table 1.**
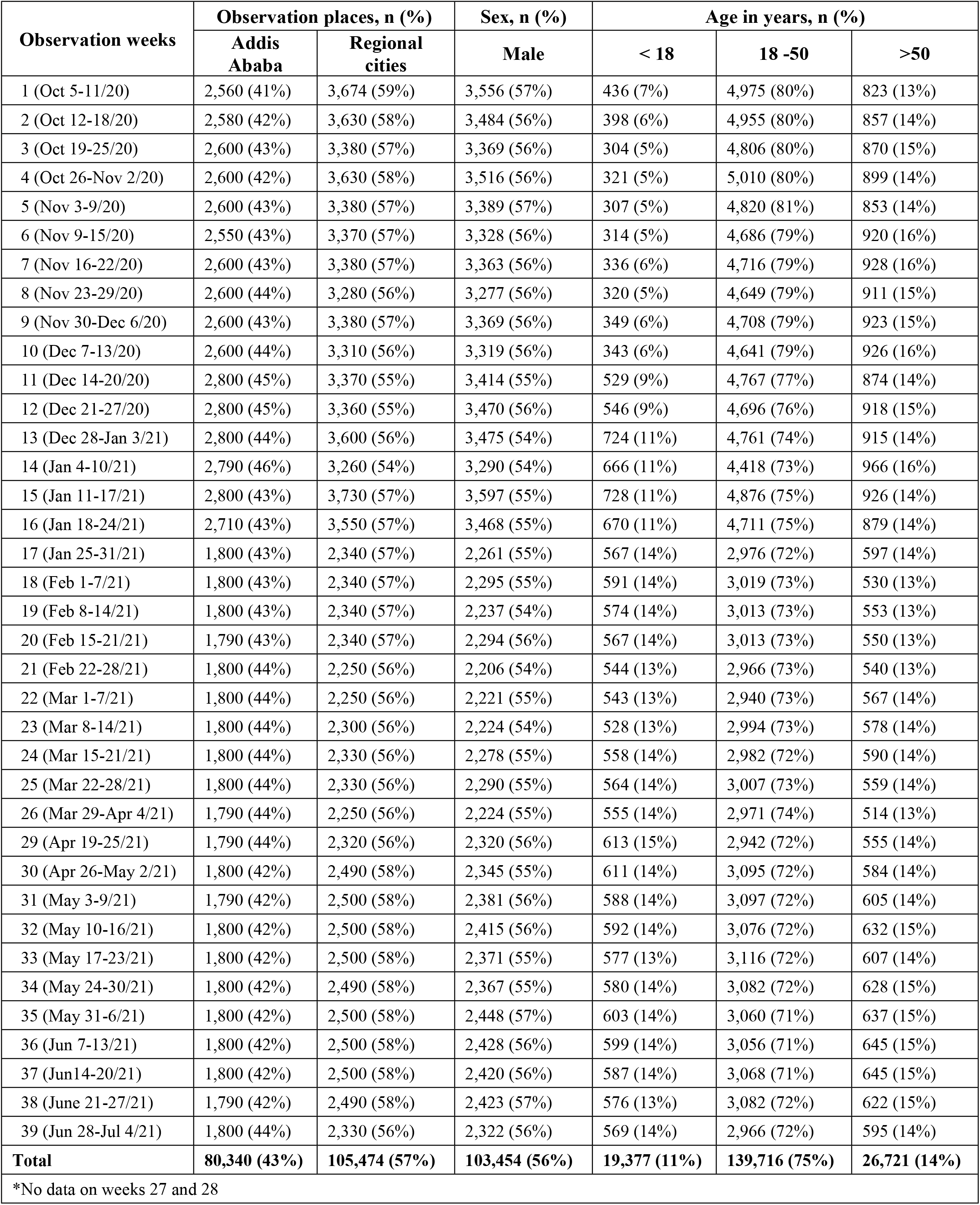
Participants observed for NPI in Ethiopia (October 2020 – July 2021)*

| Observation weeks | Observation places, n (%) |  | Sex, n (%) | Age in years, n (%) |  |  |
| --- | --- | --- | --- | --- | --- | --- |
|  | Addis Ababa | Regional cities | Male | < 18 | 18 -50 | >50 |
| 1 (Oct 5-11/20) | 2,560 (41%) | 3,674 (59%) | 3,556 (57%) | 436 (7%) | 4,975 (80%) | 823 (13%) |
| 2 (Oct 12-18/20) | 2,580 (42%) | 3,630 (58%) | 3,484 (56%) | 398 (6%) | 4,955 (80%) | 857 (14%) |
| 3 (Oct 19-25/20) | 2,600 (43%) | 3,380 (57%) | 3,369 (56%) | 304 (5%) | 4,806 (80%) | 870 (15%) |
| 4 (Oct 26-Nov 2/20) | 2,600 (42%) | 3,630 (58%) | 3,516 (56%) | 321 (5%) | 5,010 (80%) | 899 (14%) |
| 5 (Nov 3-9/20) | 2,600 (43%) | 3,380 (57%) | 3,389 (57%) | 307 (5%) | 4,820 (81%) | 853 (14%) |
| 6 (Nov 9-15/20) | 2,550 (43%) | 3,370 (57%) | 3,328 (56%) | 314 (5%) | 4,686 (79%) | 920 (16%) |
| 7 (Nov 16-22/20) | 2,600 (43%) | 3,380 (57%) | 3,363 (56%) | 336 (6%) | 4,716 (79%) | 928 (16%) |
| 8 (Nov 23-29/20) | 2,600 (44%) | 3,280 (56%) | 3,277 (56%) | 320 (5%) | 4,649 (79%) | 911 (15%) |
| 9 (Nov 30-Dec 6/20) | 2,600 (43%) | 3,380 (57%) | 3,369 (56%) | 349 (6%) | 4,708 (79%) | 923 (15%) |
| 10 (Dec 7-13/20) | 2,600 (44%) | 3,310 (56%) | 3,319 (56%) | 343 (6%) | 4,641 (79%) | 926 (16%) |
| 11 (Dec 14-20/20) | 2,800 (45%) | 3,370 (55%) | 3,414 (55%) | 529 (9%) | 4,767 (77%) | 874 (14%) |
| 12 (Dec 21-27/20) | 2,800 (45%) | 3,360 (55%) | 3,470 (56%) | 546 (9%) | 4,696 (76%) | 918 (15%) |
| 13 (Dec 28-Jan 3/21) | 2,800 (44%) | 3,600 (56%) | 3,475 (54%) | 724 (11%) | 4,761 (74%) | 915 (14%) |
| 14 (Jan 4-10/21) | 2,790 (46%) | 3,260 (54%) | 3,290 (54%) | 666 (11%) | 4,418 (73%) | 966 (16%) |
| 15 (Jan 11-17/21) | 2,800 (43%) | 3,730 (57%) | 3,597 (55%) | 728 (11%) | 4,876 (75%) | 926 (14%) |
| 16 (Jan 18-24/21) | 2,710 (43%) | 3,550 (57%) | 3,468 (55%) | 670 (11%) | 4,711 (75%) | 879 (14%) |
| 17 (Jan 25-31/21) | 1,800 (43%) | 2,340 (57%) | 2,261 (55%) | 567 (14%) | 2,976 (72%) | 597 (14%) |
| 18 (Feb 1-7/21) | 1,800 (43%) | 2,340 (57%) | 2,295 (55%) | 591 (14%) | 3,019 (73%) | 530 (13%) |
| 19 (Feb 8-14/21) | 1,800 (43%) | 2,340 (57%) | 2,237 (54%) | 574 (14%) | 3,013 (73%) | 553 (13%) |
| 20 (Feb 15-21/21) | 1,790 (43%) | 2,340 (57%) | 2,294 (56%) | 567 (14%) | 3,013 (73%) | 550 (13%) |
| 21 (Feb 22-28/21) | 1,800 (44%) | 2,250 (56%) | 2,206 (54%) | 544 (13%) | 2,966 (73%) | 540 (13%) |
| 22 (Mar 1-7/21) | 1,800 (44%) | 2,250 (56%) | 2,221 (55%) | 543 (13%) | 2,940 (73%) | 567 (14%) |
| 23 (Mar 8-14/21) | 1,800 (44%) | 2,300 (56%) | 2,224 (54%) | 528 (13%) | 2,994 (73%) | 578 (14%) |
| 24 (Mar 15-21/21) | 1,800 (44%) | 2,330 (56%) | 2,278 (55%) | 558 (14%) | 2,982 (72%) | 590 (14%) |
| 25 (Mar 22-28/21) | 1,800 (44%) | 2,330 (56%) | 2,290 (55%) | 564 (14%) | 3,007 (73%) | 559 (14%) |
| 26 (Mar 29-Apr 4/21) | 1,790 (44%) | 2,250 (56%) | 2,224 (55%) | 555 (14%) | 2,971 (74%) | 514 (13%) |
| 29 (Apr 19-25/21) | 1,790 (44%) | 2,320 (56%) | 2,320 (56%) | 613 (15%) | 2,942 (72%) | 555 (14%) |
| 30 (Apr 26-May 2/21) | 1,800 (42%) | 2,490 (58%) | 2,345 (55%) | 611 (14%) | 3,095 (72%) | 584 (14%) |
| 31 (May 3-9/21) | 1,790 (42%) | 2,500 (58%) | 2,381 (56%) | 588 (14%) | 3,097 (72%) | 605 (14%) |
| 32 (May 10-16/21) | 1,800 (42%) | 2,500 (58%) | 2,415 (56%) | 592 (14%) | 3,076 (72%) | 632 (15%) |
| 33 (May 17-23/21) | 1,800 (42%) | 2,500 (58%) | 2,371 (55%) | 577 (13%) | 3,116 (72%) | 607 (14%) |
| 34 (May 24-30/21) | 1,800 (42%) | 2,490 (58%) | 2,367 (55%) | 580 (14%) | 3,082 (72%) | 628 (15%) |
| 35 (May 31-6/21) | 1,800 (42%) | 2,500 (58%) | 2,448 (57%) | 603 (14%) | 3,060 (71%) | 637 (15%) |
| 36 (Jun 7-13/21) | 1,800 (42%) | 2,500 (58%) | 2,428 (56%) | 599 (14%) | 3,056 (71%) | 645 (15%) |
| 37 (Jun 14-20/21) | 1,800 (42%) | 2,500 (58%) | 2,420 (56%) | 587 (14%) | 3,068 (71%) | 645 (15%) |
| 38 (Jun 21-27/21) | 1,790 (42%) | 2,490 (58%) | 2,423 (57%) | 576 (13%) | 3,082 (72%) | 622 (15%) |
| 39 (Jun 28-Jul 4/21) | 1,800 (44%) | 2,330 (56%) | 2,322 (56%) | 569 (14%) | 2,966 (72%) | 595 (14%) |
| **Total** | **80,340 (43%)** | **105,474 (57%)** | **103,454 (56%)** | **19,377 (11%)** | **139,716 (75%)** | **26,721 (14%)** |
| *No data on weeks 27 and 28 |  |  |  |  |  |  |

### Overall trends of Proper NPI practice

#### Proper respiratory hygiene

From the three NPIs, the community had better compliance to respiratory hygiene followed by physical distance than hand hygiene. The respiratory hygiene compliance at a national level was 41% at the start of the observation week and declined to the lowest proportion of 32% holding for several weeks, then showed weekly progress up to 48% and maintained through the 26^th^ to the 30^th^ weeks Again, the proportion of proper respiratory hygiene compliance showed a continuous decline to 39% until the last observation week (Figure 2). However, there was a big difference in respiratory hygiene compliance between Addis Ababa city and major regional cities; there was higher compliance in Addis Ababa city across the observation period than in the regional cities and this difference is statistically significant at P<0.001. Increased compliance was also observed in both Addis Ababa and regional cities around the 26^th^through the 30^th^ weeks (Figure 3)

**Figure 2.**
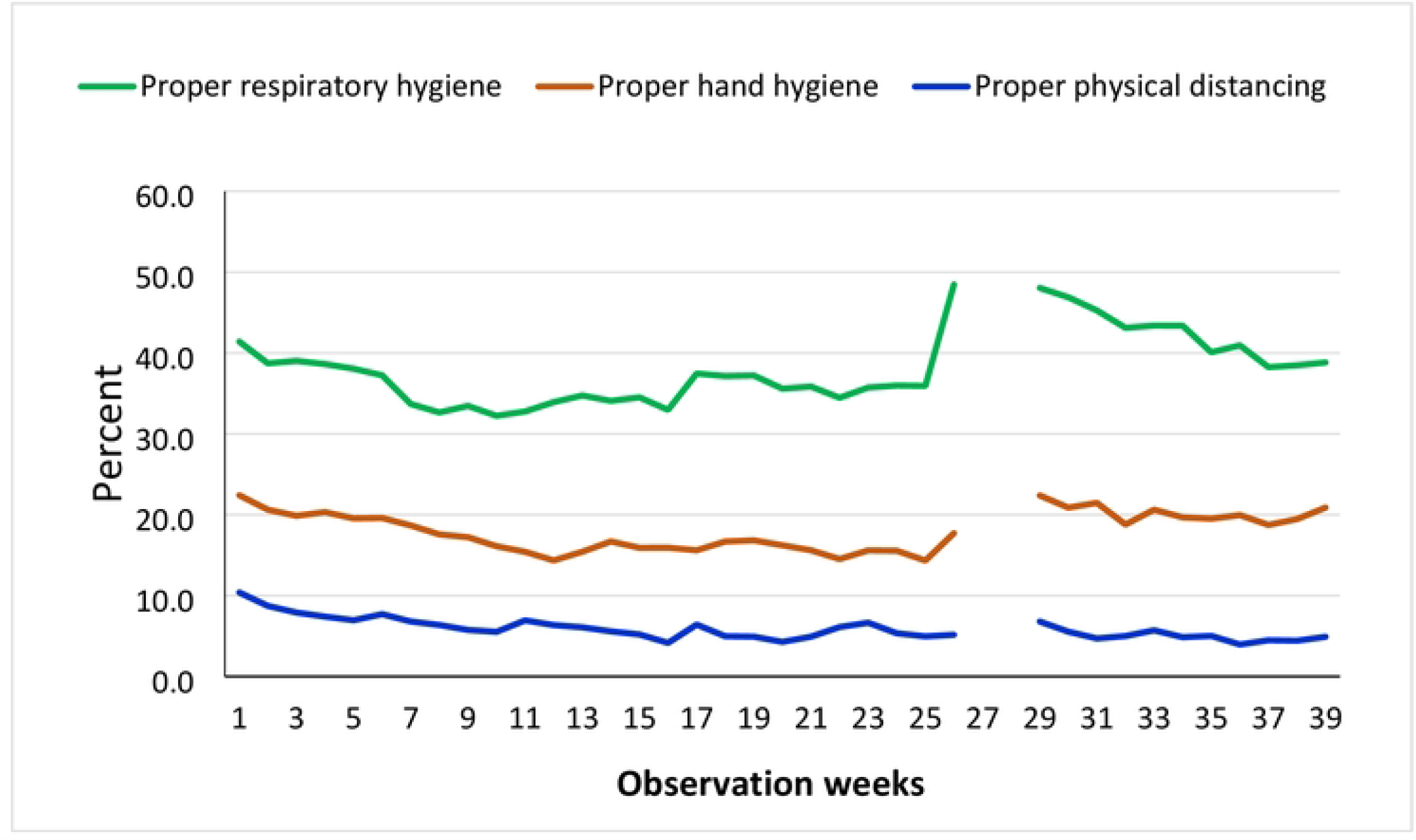
Overall city-wide trends of proper NPI practices for 39 weeks, Oct 5/2020-July 4/2021. **Note:** Data were not collected on week 27 and 28

**Figure 3.**
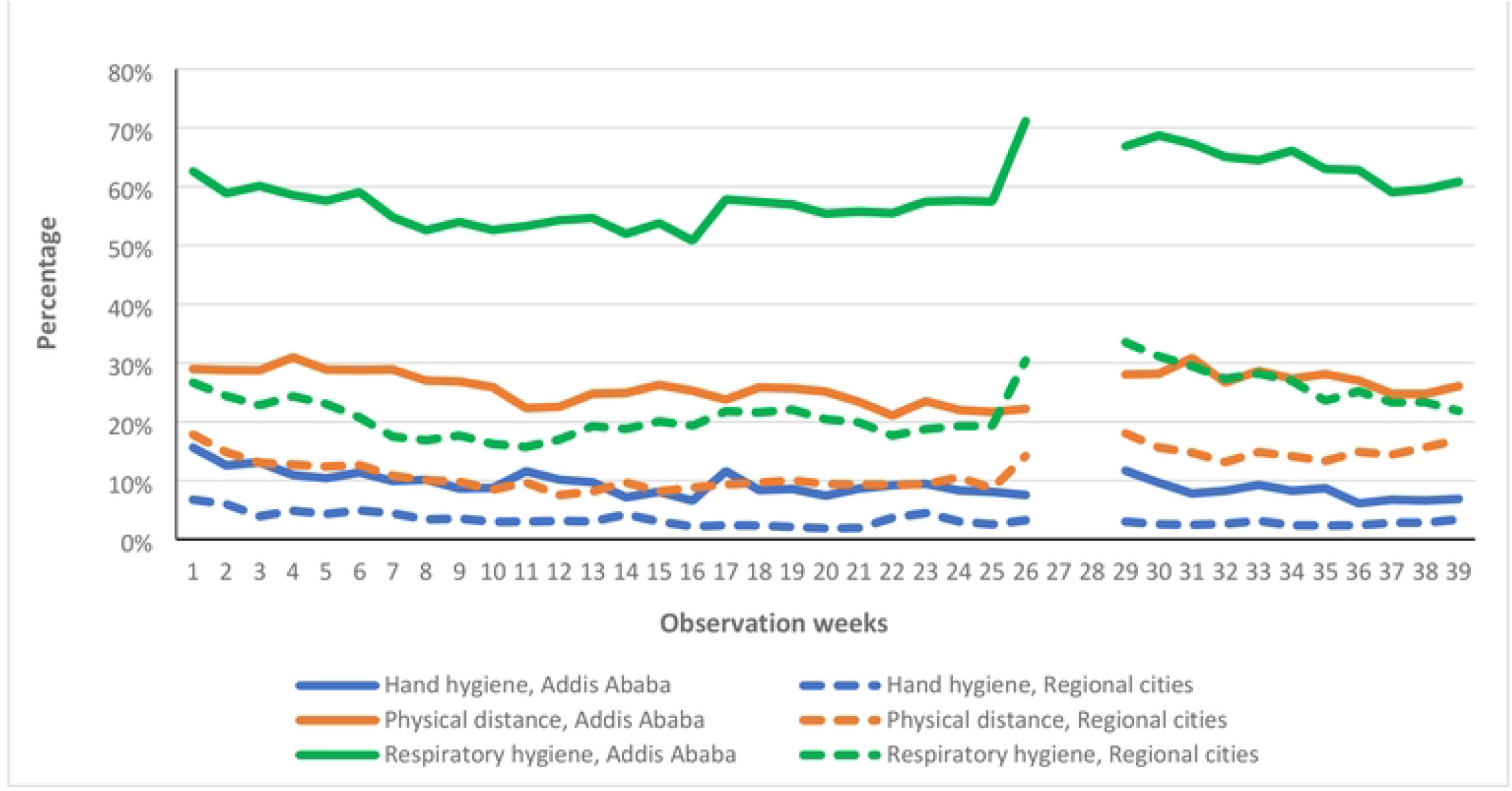
Trends of proper NPI practice in the Addis Ababa vs regional cities, Oct 5/2020-July 4/2021.

#### Proper physical distancing

Proper physical distancing was the second NPI practice the community complied with next to respiratory hygiene. The proportion of proper physical distancing at a national level range from 14% - 22% in the observation weeks. Similar to respiratory hygiene, the declined weekly trend from the start showed a small increment around the 29^th^through the 31^st^ weeks (Figure 2). The proportion of proper physical distancing across the observation period in Addis Ababa ranged from 21% to 31% whereas the proportion in major regional cities was 8% - 18 %. There is a big difference in proper physical distance practice between Addis Ababa city and regional cities, this difference is again statistically significant at P<0.001. (Figure 3).

#### Proper hand Hygiene

Proper hand hygiene was the least NPI practice the community complied. The proportion at national level ranges from 4% - 10% during the observation weeks (Figure 2). The overall proportion of proper hand hygiene practice showed a declining trend in both Addis Ababa city and major regional cities. However, there was still a big difference in the proportion of proper hand hygiene between the two population groups, ranges from 7% - 16% and 2% - 7%, respectively. This difference is also statistically significant at P<0.001 (Figure 3).

### Variation of proper NPI practice in service facilities

#### Proper respiratory hygiene

An overall analysis combining the 39 weeks observation data showed that the proportion of proper respiratory hygiene compliance at the different service facilities at national level ranges from 25% - 54%; the highest compliance was observed at the bank and the least at food and drink establishments. The stratified analysis of proper respiratory hygiene by cities for the different facilities ranged from 40% - 80% and 14% - 35% in Addis Ababa and regional cities, respectively. Highest public compliance was observed in Addis Ababa at all the service facilities compared to the regional cities (P<0.001) (Figure 4).

**Figure 4.**
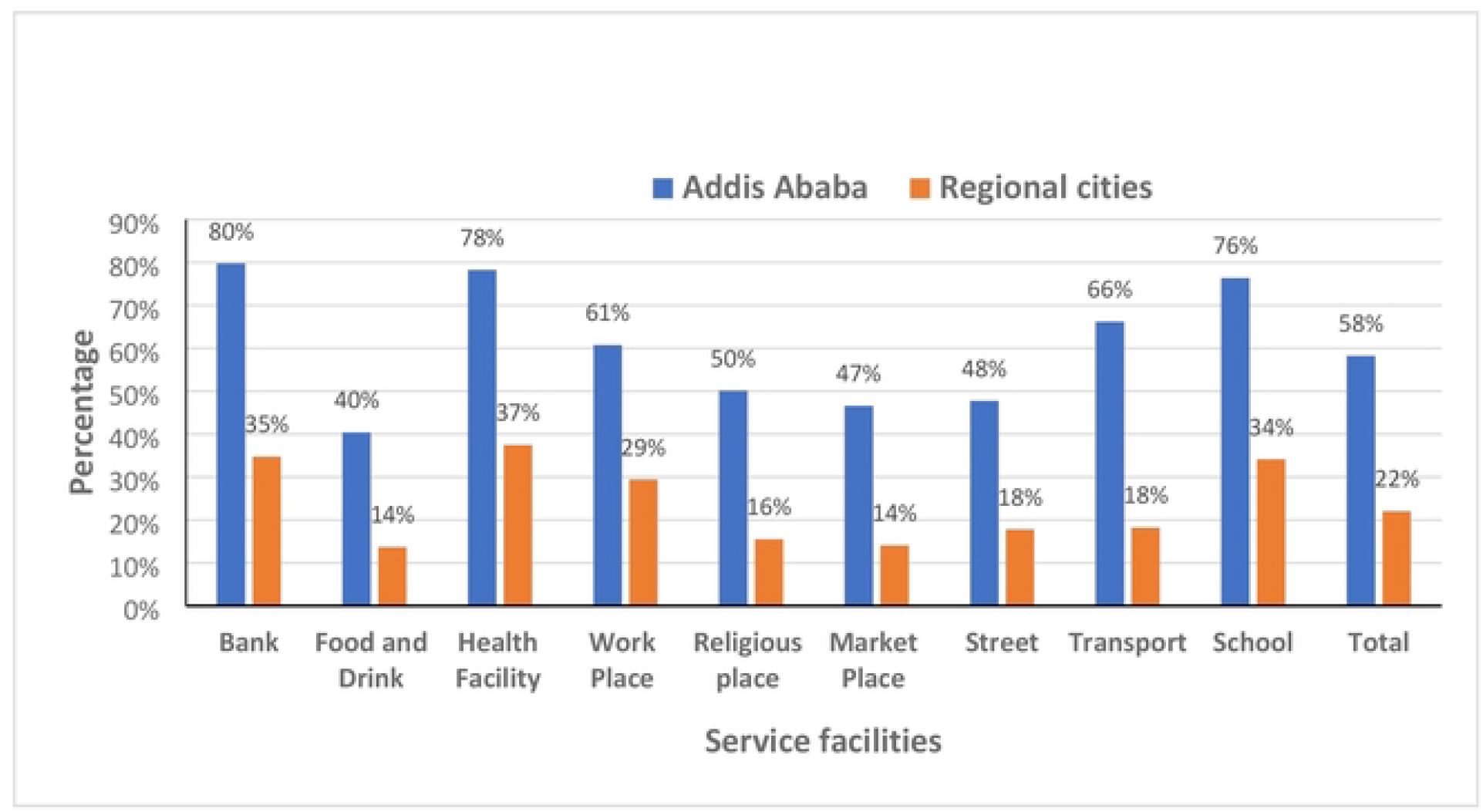
Proper respiratory hygiene at different service facilities in capital Vs regional cities, Oct 5/2020-July 4/2021.

#### Proper physical distancing

The proper physical distancing public compliance at the different service facilities at the national level ranges from 9% - 23%. Similar to the respiratory hygiene the highest compliance was recorded at the bank but the least at the transport service. The stratified analysis of proper physical distancing by observation places for the different facilities ranges from 13% - 35% and 7% - 16% in Addis Ababa and regional cities, respectively. The highest and the least proportion of compliance in Addis Ababa was at the bank and transport service facilities, respectively. Although the least proportion of compliance in regional cities was at the transport service but the highest compliance was at the workplace. An overall highest public compliance was observed in Addis Ababa at all the service facilities compared to the regional cities. This difference is statistically significant at P<0.001 (Figure 5).

**Figure 5.**
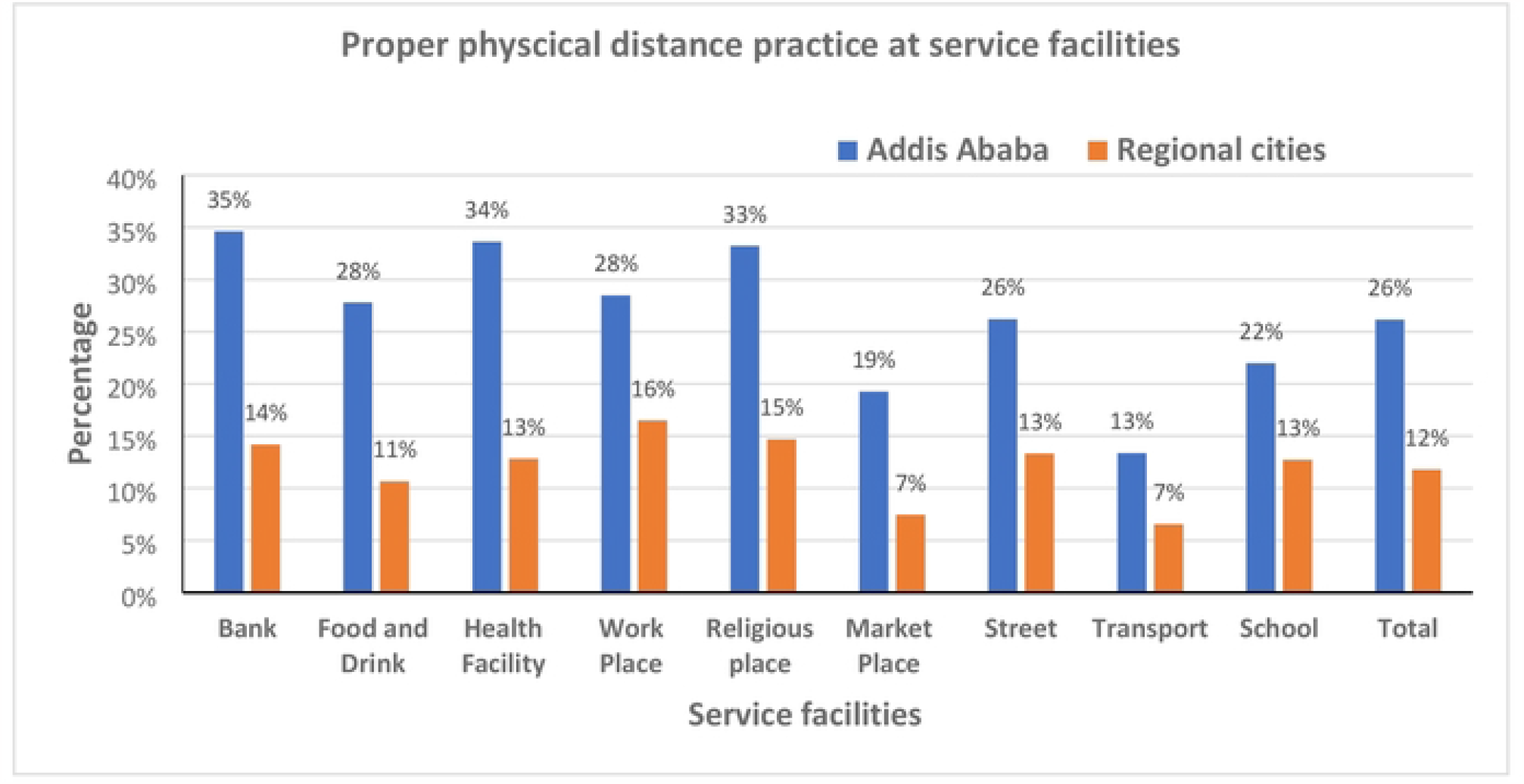
The proper physical distancing at different service facilities in capital vs regional cities, Oct 5/2020-July 4/2021.

#### Proper hand hygiene

The proper hand hygiene public compliance at the different service facilities at the national level ranges from 0.2% - 16%. The highest compliance was recorded at the food and drink establishments but the least at the transport service. The stratified analysis of proper hand hygiene by cities for the different facilities ranges from 0.3% - 20% and 0.1% - 12% in Addis Ababa and regional cities, respectively. Similar to the national level, the highest and the least proportion of compliance was recorded at the same facilities in both cities. An overall highest public compliance was observed in Addis Ababa city at all the service facilities compared to the regional cities, this difference is statistically significant at P<0.001 (Figure 6).

**Figure 6.**
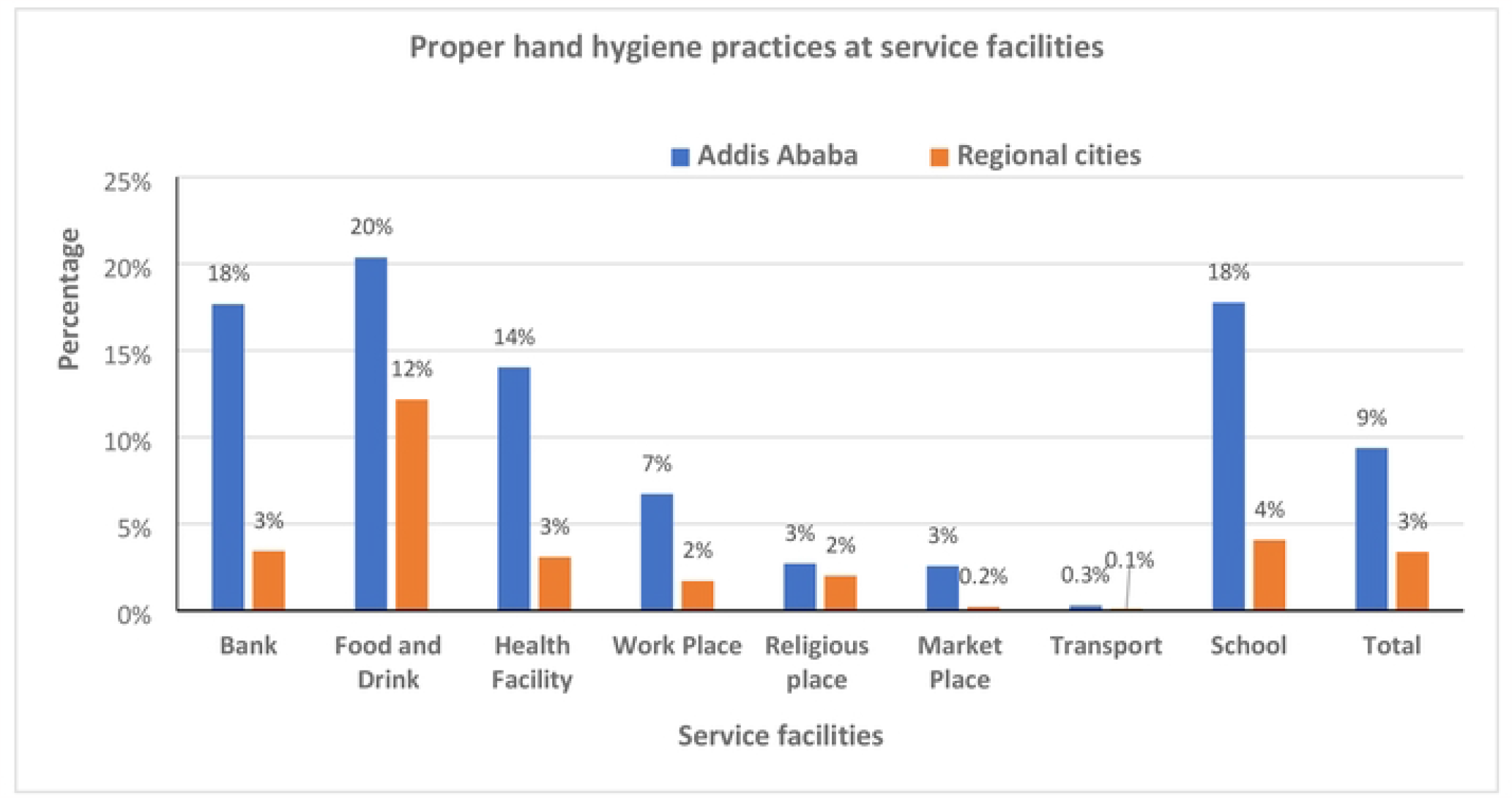
Proper hand hygiene at different service facilities in capital Vs regional cities, Oct 5/2020-July 4/2021.

### Variation of proper NPI practice by sex and age

A stratified analysis was performed to observe NPI practice difference by sex and age group within the observation city. The practice of the three NPIs in Addis Ababa was more than that in the regional cities, with statistical differences (p<0.001). Females tend to have increased respiratory hygiene practice relative to males, while the age group greater than 50 had better physical distancing than other age groups (Table 2).

**Table 2.** Proper NPI practices by sex and age group in the capital Vs regional cities, Oct 5/2020-July 4/2021.

| NPI practices | Observation Cities | Sex |  |  | Age, years |  |  | P-value |
| --- | --- | --- | --- | --- | --- | --- | --- | --- |
|  |  | Male, n (%) | Female, n (%) | P-value | <18, n (%) | 18-50, n (%) | >50, n (%) |  |
| **Proper respiratory hygiene** | Addis Ababa | 23,264 (55%) | 23,510 (62%) | <0.001 | 5,183 (64%) | 35,586 (58%) | 6,005 (57%) | <0.001 |
|  | Regional cities | 12,968 (21%) | 10,257 (23%) | <0.001 | 2,390 (21%) | 16,951 (22%) | 3,884 (24%) | <0.001 |
| **Proper physical distancing** | Addis Ababa | 11,747 (28%) | 9,276 (24%) | <0.001 | 1,620 (20%) | 16,171 (26%) | 3,232 (31%) | <0.001 |
|  | Regional cities | 7,539 (12%) | 4,887 (11%) | <0.001 | 1,036 (9%) | 9,067 (12%) | 2,323 (14%) | <0.001 |
| **Proper hand hygiene** | Addis Ababa | 3,416 (8%) | 3,144 (8%) | 0.267 | 1,089 (13%) | 4,723 (8%) | 748 (7%) | <0.001 |
|  | Regional cities | 2,064 (3%) | 1,030 (2%) | <0.001 | 259 (2%) | 2,433 (3%) | 402 (3%) | <0.001 |

## Discussion

The overall trend of NPI practice in the community showed a decline trend for the last ten weeks. Respiratory hygiene is the most widely practiced NPI by individuals than the other two NPIs both in Addis Ababa and regional cities. Generally, all the three NPIs are more practiced in Addis Ababa than regional cities. Besides, the highest NPI practice was observed at the formally organized service facilities such as workplaces and banks. In addition, there was NPI compliance level difference in the community by their sociodemographic characteristics.

The overall trends of the three NPIs are slightly declining specially on wards around the 31^st^ week after all the NPIs have reached on peak during the weeks of 26^th^ – 30^th^. For the rest of the observation period, for 25 weeks prior to the peak, there was no significant change in the level of NPI compliance. An increased community compliance level started at the 26^th^ week (the last week of march) for all the three NPIs, but the rate of change from the previous week is quite different for each NPI; highest comply level to respiratory hygiene from 36% to 49% and least comply to hand hygiene from 5.0% to 5.2%. The increasing community compliance level during these weeks might be linked with several factors. One of the main factors might be the trends of Covid-19 prevention public measures that has been taken during this period. Every government world-wide have taken different types of public measures at different time to reduce the spread of Covid-19 infection[1, 16-18]. During the 39 weeks of observation period, a total of 76 WHO documented Covid-19 prevention measures were implemented in Ethiopia. From these new interventions 13 of them were implemented during four weeks period in the month of March. The intensity of public measure implementation programs in the month of march was highest on average 3-4 new interventions per week, whereas in the rest of the months 1-2 interventions per week. Besides, several of the measures were related to the mandatory wearing of face mask in public places followed by physical distancing. Although there were new additional interventions related to general public awareness, policy and regulation programs but there was no specific to hand hygiene related intervention programs initiated during this period. [1]. Therefore, the trends and the type of risk communication public measures during the month of march might be the source of ignition for the highest overall NPI compliance level during the 26^th^ – 30^th^ weeks.

Most of NPI related studies focused on the effectiveness of different and combined NPIs on the control of transmission but information regarding public compliance level with time-line was limited. In some studies, organized efforts initiated by governments, public awareness advocacy and risk communication programs affect NPI adherence in a community [19-21]. A population based bi-weekly survey (April to November 2020) from the US showed a substantial decrease of NPI adherence index from 70 at the beginning of the survey, maintain plateau at 50s during June and a slight increase to 60 in November [21]. The current author also reported a similar pattern of change in NPI compliance level in the community [11]. Another community-based study that examine covid-19 community mitigation practices in Nigeria found that wide ranges of government measures, healthcare policy related to the pandemic and public campaign intervention programs affected individual behavior towards NPIs practices [20].

Throughout the observation period, proper respiratory hygiene compliance is consistently the highest from the three NPI practices in our study. At the beginning of the pandemic social distancing and handwashing were well accepted NPIs by many people worldwide as the most effective strategy to control the transmission compared to the face mask [22]. Even before changing the current guideline WHO also declared as there was no evidence about face mask in protecting healthy individuals from Covid-19 infection [23]. However, the embarking message by CDC about the benefit of face covering for the prevention of transmission against WHO influence people practice towards face mask use [24]. Lately, the prevention of Covid-19 transmission through proper respiratory hygiene (wearing mask) is widely advocated throughout the world. On top of the regulatory public measures, several scientific evidences from the health sciences, advices from recognized public health institutions promote face covering as an efficient prevention method specially in resource limited settings [24-27]. Public availability of this information and the increased rate of Covid 19 transmission might influence evidence based informed decision-making power of individuals to comply to face mask than the other NPIs. The feasibility of enforcing the mandatory use of face-mask at individual level while in service facilities and public places might also a reason for the higher proper respiratory hygiene compliance than other NPIs in the current study [4].

The level of compliance to each NPI is affected by several factors. The current study highlighted the variation of NPI compliance of individuals by place of residence, socio-demographic characteristics and at service provision facilities. People living in the capital city had a better compliance to the overall NPIs compared to regional cities residents. Except on physical distancing females were better compliance to the other NPIs than males and people at the bank and workplaces have better NPI compliance than at the transport service provision. Abdelhafiz and colleagues identified urban resident Egyptians had better knowledge and NPI practice compared to the rural residents [28]. Females were more likely to follow good preventive practices, comply with wearing mask and hand hygiene than men in different population-based studies in Africa [11, 20, 29]. A previous weekly monitoring of NPIs at different service facilities have also a similar result [11]. The availability of information, restriction and required procedures at institutions might be the main reason people to comply with NPI measures.

Although monitoring weekly NPI practice data from the community is an important source of information for decision, but this study has also several limitations which was mentioned in previous publication [11].

## Conclusions and recommendations

Generally, community compliance to NPI practice showed a decline trend in Ethiopia but an increased compliance was also observed following the implementation of government initiated public measures. Comply to proper respiratory hygiene is by far higher than hand hygiene and physical distancing. Besides, the overall NPI practice level is varied by place of resident, sociodemographic characteristics of individuals and service provision facilities. Therefore, public initiated risk communication and advocacy measures should be strengthened to increase community compliance to NPI practices.

## Data Availability

In the manuscript and additional SPSS data file uploaded

## Acknowledgments

We would like to thank the Federal Democratic Republic of Ethiopia Ministry of Health and the College of Health Sciences and the School of Public Health of Addis Ababa University for supporting this study.

## Author’s contributions

**Conceptualization**: Damen Hailemariam, Abera Kumie, Samson Wakuma, Yifokire Tefera.

**Data curation**: Abera Kumie, Yifokire Tefera, Shibabaw Yirsaw.

**Formal analysis**: Abera Kumie, Yifokire Tefera, Samson Wakuma.

**Methodology**: Abera Kumie, Samson Wakuma, Yifokire Tefera, Teferai Abegaz.

**Software**: Abera Kumie, Yifokire Tefera, Shibabaw Yirsaw.

**Supervision**: Damen Hailemariam, Abera Kumie, Yifokire Tefera, Samson Wakuma, Teferi Abegaz.

**Validation**: Abera Kumie, Yifokire Tefera, Teferi Abegaz.

**Visualization**: Damen Hailemariam, Yifokire Tefera, Abera Kumie

**Writing** – original draft: Abera Kumie, Yifokire Tefera, Teferi Abegaz, Samson Wakuma.

**Writing** – review & editing: Damen Hailemariam, Abera Kumie, Samson Wakuma, Yifokire Tefera, Teferi Abegaz.

## Reference

1. World Health Organization, Tracking Public Health and Social Measures A Global Dataset. World Health Organization, 2020.

2. Haug, N., et al., Ranking the effectiveness of worldwide COVID-19 government interventions. Nature human behaviour, 2020. 4(12): p. 1303–1312.

3. Chan, L.Y.H., B. Yuan, and M. Convertino, COVID-19 non-pharmaceutical intervention portfolio effectiveness and risk communication predominance. Scientific reports, 2021. 11(1): p. 1–17.

4. Brauner, J.M., et al., The effectiveness of eight nonpharmaceutical interventions against COVID-19 in 41 countries. MedRxiv, 2020.

5. Anderson, R.M., et al., How will country-based mitigation measures influence the course of the COVID-19 epidemic? The lancet, 2020. 395(10228): p. 931–934.

6. World Health Organization, COVID-19 global risk communication and community engagement strategy, December 2020-May 2021: interim guidance, 23 December 2020. 2020, World Health Organization.

7. Sharma, M., et al., How robust are the estimated effects of nonpharmaceutical interventions against COVID-19? Advances in Neural Information Processing Systems, 2020. 33: p. 12175–12186.

8. Rowan, N.J. and R.A. Moral, Disposable face masks and reusable face coverings as non-pharmaceutical interventions (NPIs) to prevent transmission of SARS-CoV-2 variants that cause coronavirus disease (COVID-19): Role of new sustainable NPI design innovations and predictive mathematical modelling. Science of the Total Environment, 2021. 772: p. 145530.

9. Ethiopian Public Health Institute, Ethiopia Covid-19 Monitoring platform. https://www.covid19.et/covid-19/. 2022.

10. Ethiopian Public Health Institute, Covid-19 Guidelines. https://covid19.ephi.gov.et/covid-19/. 2022.

11. Hailemariam, D., et al., Trends in non-pharmaceutical intervention (NPI) related community practice for the prevention of COVID-19 in Addis Ababa, Ethiopia. PloS one, 2021. 16(11): p. e0259229.

12. Deressa, W., et al., Risk perceptions and preventive practices of COVID-19 among healthcare professionals in public hospitals in Addis Ababa, Ethiopia. PLoS One, 2021. 16(6): p. e0242471.

13. Federal Negarit Gazett of the Federal Democratic Republic of Ethiopia, Regulation No.466/2020 State of Emergency Proclamation No. 3/2020. https://covidlawlab.org/wp-content/uploads/2021/07/State-of-Emergency-Proclamation-No.-3-2020-Implementation-Regulation-Ethiopia.pdf. 2020.

14. Fikrie, A., E. Amaje, and W. Golicha, Social distancing practice and associated factors in response to COVID-19 pandemic at West Guji Zone, Southern Ethiopia, 2021: A community based cross-sectional study. PloS one, 2021. 16(12): p. e0261065.

15. Ethiopian Statistic Service, Central Statical Agency. Population Projections for Ethiopia 2007-2037. https://www.statsethiopia.gov.et/population-projection/. 2013.

16. Chowdhury, R., et al., The Global Dynamic Interventions Strategies for COVID-19 Collaborative Group. Dynamic interventions to control covid-19 pandemic: a multivariate prediction modelling study comparing 16 worldwide countries. Eur J Epidemiol, 2020. 35(5): p. 389–399.

17. Lai, S., et al., Effect of non-pharmaceutical interventions to contain COVID-19 in China. nature, 2020. 585(7825): p. 410–413.

18. Flaxman, S., et al., Estimating the effects of non-pharmaceutical interventions on COVID-19 in Europe. Nature, 2020. 584(7820): p. 257–261.

19. Dan-Nwafor, C., et al., Nigeria’s public health response to the COVID-19 pandemic: January to May 2020. Journal of Global Health, 2020. 10(2).

20. Shittu, E., et al. Examining Psychosocial Factors and Community Mitigation Practices to Limit the Spread of COVID-19: Evidence from Nigeria. in Healthcare. 2022. MDPI.

21. Crane, M.A., et al., Change in reported adherence to nonpharmaceutical interventions during the COVID-19 pandemic, April-November 2020. Jama, 2021. 325(9): p. 883–885.

22. Zhang, L., et al., Can self-imposed prevention measures mitigate the COVID-19 epidemic? PLoS medicine, 2020. 17(7): p. e1003240.

23. World Health, O., Advice on the use of masks in the community, during home care and in health care settings in the context of the novel coronavirus (2019-nCoV) outbreak: interim guidance, 29 January 2020. 2020, World Health Organization: Geneva.

24. CDC, Center for Disease Control and Prevention. Recommendation Regarding the Use of Cloth Face Covering. https://www.cdc.gov/coro. 2020.

25. Li, T., et al., Mask or no mask for COVID-19: A public health and market study. PloS one, 2020. 15(8): p. e0237691.

26. World Health, O., Advice on the use of masks in the context of COVID-19: interim guidance, 6 April 2020. 2020, World Health Organization: Geneva.

27. Ethiopian Public Health Institute, Directive No 30/2020 A DIRECTIVE ISSUED FOR THE PREVENTION AND CONTROL OF COVID-19 PANDEMIC. https://covid19.ephi.gov.et/wp-content/uploads/2020/10/Registerd-COVID-19-Directive-2013_Final_051020.pdf. 2020.

28. Abdelhafiz, A.S., et al., Knowledge, perceptions, and attitude of Egyptians towards the novel coronavirus disease (COVID-19). Journal of community health, 2020. 45(5): p. 881–890.

29. Nwonwu, E.U., et al., Knowledge and preventive practice to COVID-19 among household heads in Enugu metropolis, South-East Nigeria. The Pan African Medical Journal, 2020. 37.

